# Computational characterization of inhaled droplet transport in the upper airway leading to SARS-CoV-2 infection

**DOI:** 10.1101/2020.07.27.20162362

**Authors:** Saikat Basu

## Abstract

How human respiratory physiology and the transport phenomena associated with the inhaled airflow therein proceed to impact transmission of SARS-CoV-2, leading to the initial infection, is an open question. An answer can help determine the susceptibility of an individual on exposure to a COVID-2019 carrier and can also quantify the still-unknown infectious dose for the disease. Synergizing computational fluid mechanics enabled tracking of respiratory transport in medical imaging-based anatomic domains, with sputum assessment data from hospitalized COVID-19 patients and earlier measurements of ejecta size distribution during regular speech – this study shows that the regional deposition of virus-laden inhaled droplets at the initial nasopharyngeal infection sites peaks for the droplet size range of 2.5 – 19 microns, and reveals that the number of virions that go on to establish the infection can be merely in the order of hundreds.

## Introduction

Severe acute respiratory syndrome coronavirus 2 (SARS-CoV-2) has been identified as the causative agent for coronavirus disease 2019 (COVID-19), that has inflicted a global pandemic with nearly 34 million confirmed infections and over 1 million deaths worldwide, as of late-September 2020; for details, see^1^.

As is well-known by now, transmission of respiratory infections such as COVID-19 occurs through carriage of pathogens via droplets of different sizes produced during sneezing, coughing, singing, normal speech, and even, breathing^2^. Accordingly, the means of person-to-person infection are projected to be three-way^3^: (a) inhalation of virus-laden droplets emitted by an infected individual at close-range; (b) inhalation of vaporized droplet nuclei that can float in air for hours; and (c) contaminating the respiratory mucosa through physical contact to external surfaces (*fomites*) with droplet deposits sitting on them. While (a) is valid for short-distance exposures to the COVID-19 carrier, transmission through modes (b) and (c) can happen over larger distances and longer time scales. However, clustering trends of infection spread (e.g. in industrial units^4^, in closed groups^5^, and inside households^6^) suggest that close-range exposures can be a critical determinant in worsening the pandemic. A follow-up question might be – *what entails an exposure?* A key component therein are the respiratory droplet sizes one is exposed to. Coughing and sneezing typically generate droplets with length-scales of *𝒪* (10^2^) to *𝒪* (10^3^) *µ*, while oral droplets ejected during normal speaking can range over *∼*0.1*−*500 *µ*^3,7^. The main competing effects determining the fate of these droplets are the ambient temperature and humidity (e.g. low relative humidity induces fast evaporation and shrinkage of the droplets), and the size of the droplet that controls its inertia and the gravitational force acting on it. While smaller droplets would stay airborne for longer, the larger droplets tend to fall fast ballistically; with the critical size for this transition being in the vicinity of 100 *µ*^8, 9^. Of note here, this study does not insist on any nomenclatural distinction between “aerosols” and “droplets” owing to ambiguities^10^ in common perception, and simply refers to all expiratory liquid particulates as *droplets*.

For tracking what range of virus-bearing droplet sizes might be more potent for SARS-CoV-2 transmission and to eventually induce infection, it is key that we identify the initial infection sites. At least two recent studies^11,12^ reveal a striking pattern of relatively high SARS-CoV-2 infectivity in ciliated epithelial cells along the nasal passage lining in the upper airway, to less infectivity in cells lining the throat and bronchia, and finally to relatively low infectivity at the lung cells. The trend is decidedly governed through angiotensin-converting enzyme 2 (ACE2), which is a single-pass type I membrane protein and is the surface receptor that the virus utilizes to intrude into cells. ACE2 is abundant on ciliated epithelial cells, but is relatively scarce on the surface of the lower airway cells. While these findings are for *in vitro* samples, deposition of virus-laden droplets along the anterior nasal airway might not be so effective as to launch an infection despite the presence of ciliated cells, since the mucus layer provides some protection against virus invasion and infection^3^. This sets up *nasopharynx* (i.e. the region in the upper airway posterior to the septum and comprising the superior portion of the pharynx; for reference, see Figure 1, Panel A) as the main *initial* infection site; it acts as the seeding zone for subsequent infection of the lower airway via aspiration of virus-laden boluses of nasopharyngeal fluids. The *ansatz* is supported by the efficacy^13^ of nasopharyngeal swab testing for COVID-19 diagnosis, when compared to oropharyngeal swabs. So at this point, a valid question to ask would be: *what are the dominant inhaled droplet sizes that are making their way to the nasopharynx?*

**Figure 1.**
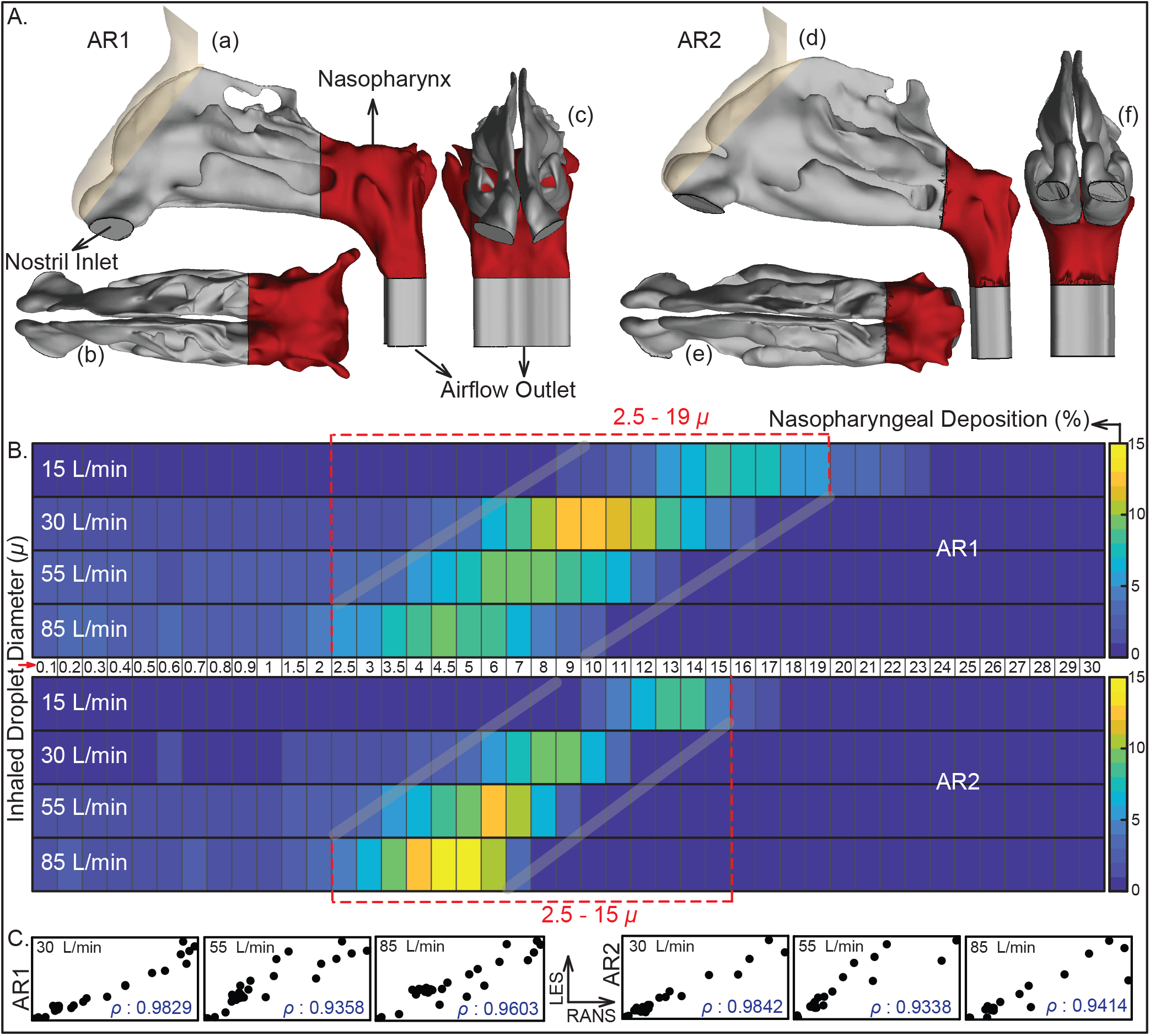
Anatomically realistic nasal geometries and the computed transport trends therein: **A**. The sagittal, axial, and coronal views of the CT-based nasal domains, shown respectively in a–c for anatomic reconstruction 1 (AR1) and d–f for anatomic reconstruction 2 (AR2). Nasopharynx is marked in red. **B**. Visuals of heat-maps for inspiratory transmission trends, showing the percentage of droplets of each size undergoing nasopharyngeal deposition (NPD). Data for different inhaled airflow rates are arranged along separate rows. Tracked droplet sizes are along the horizontal axis (positioned between the two heat-maps for AR1 and AR2). NPD peaks for droplets sized between 2.5 – 19 *µ* in AR1 and 2.5 – 15 *µ* in AR2. **C**. The correlation between RANS-based SST k-*ω* and LES results for the higher airflow rates i.e. 30, 55, and 85 L/min; therein the first three frames (bottom-left) are for AR1, the other three frames (bottom-right) correspond to data for AR2. The frames are on an aspect ratio of 0.5; *ρ* represents the Pearson’s correlation coefficient.

Respiratory droplets, on being expelled, typically lose water and shrink; – the extent of which partially depends on the fraction of non-volatile constituents present in the droplets, e.g. dehydrated epithelial cell remnants, white blood cells, enzymes, DNA, sugars, electrolytes etc. So, although sputum is composed of 99.5% water; ejected droplets, on dehydration, have a higher density of 1.3 g/ml^14^, which is what has been used for droplet tracking simulations here. This considers that the non-volatile weight fraction is in the 1 – 5% range. Such dehydration contracts the expelled droplet diameter to 27 – 34% of the initial size. Thus, for a mean 30% shrinkage and considering 100 *µ* as the critical size prompting ballistic sedimentation, this study tracks inhaled droplet sizes in the range of 0.1 *µ* to 30% of 100 *µ*, i.e. 30 *µ*. Choice of the smallest tracked droplet size is dictated by SARS-CoV-2 dimension, which is in between 0.08 – 0.2 *µ*, with an average physical diameter of 0.1 *µ*^15^.

Next piece in this puzzle relates to the breathing parameters. Allometric relations^16^ put the minute inhalation at 18.20 L/min for a 75-kg male and 15.05 L/min for a 75-kg female, for gentle steady breathing while sitting awake. In general, inspiratory rates can stretch over *∼* 15 – 85 L/min, based on whether the individual is inhaling gently or breathing in forcefully. This study simulates droplet transmission at four different inhalation rates, viz. 15, 30, 55, and 85 L/min; notably these discrete flow rates are the ones traditionally used^17^ for checking filtration capacities of protective face-coverings and respirators. The flow physics undergo a transition over this range; e.g. 15 L/min through nasal conduits is in laminar regime, the transport mechanism however devolves into turbulence at higher inhalation rates.

## Methods

### Anatomic geometry reconstruction

Computed tomography (CT)-based *in silico* model generation was accomplished according to relevant guidelines and regulations, with the anatomic geometries being reconstructed from existing de-identified imaging data from two CT-normal subjects. The use of the archived and anonymized medical records was approved with exempt status by the Institutional Review Board of the University of North Carolina (UNC) at Chapel Hill, with the requirement of informed consent being waived for retrospective use of the de-identified scans in computational research. The test subjects include a 61 year-old female (subject for anatomic reconstruction 1, or AR1) and a 37 year-old female (subject for anatomic reconstruction 2, or AR2). In context to the imaging resolution, the CT slices were collected at coronal depth increments of *∼* 0.4 mm. The nasal airspaces were extracted from the medical grade scans over a delineation range of -1024 to -300 Hounsfield units, and was complemented by careful hand-editing of the selected pixels to ensure anatomic accuracy. For this step, the DICOM (Digital Imaging and Communications in Medicine) scans for each subject were imported to the image processing software Mimics 18.0 (Materialise, Plymouth, Michigan).

The reconstructed geometries were imported as stereolithography files to ICEM-CFD 15.0 (ANSYS, Inc., Canonsburg, Pennsylvania), and then meshed spatially into minute volume elements. Conforming with established mesh refinement-based protocols^18,19^, each computational grid contained more than 4 million unstructured, graded tetrahedral elements (e.g. 4.54 million in AR1, 4.89 million in AR2); along with three prism layers of 0.1-mm thickness at the airway walls, with a height ratio of 1. The nostril inlet planes comprised 3015 elements in AR1 (1395 elements on left nostril plane, 1620 elements on right nostril plane) and 3000 elements in AR2 (1605 on left nostril plane, 1395 on right nostril plane).

### Numerical simulations

The study considers droplet transport for four different inhaled airflow rates, viz. 15, 30, 55, and 85 L/min. The lower flow rate (i.e. 15 L/min) corresponds to comfortable resting breathing, with the viscous-laminar steady-state flow physics model standing in as a close approximation^20–31^. At higher flow rates (extreme values of which may sometimes lead to nasal valve collapse), the shear layer separation from the tortuous walls of the anatomic geometries results in turbulence^32–35^. While accounting for the turbulent characteristics of the ambient airflow, the study averages the droplet deposition percentages from implementation of two distinct categories of numerical schemes, viz. (a) shear stress transport (SST) based k-*ω* model, which is a sub-class under Reynolds-averaged Navier Stokes (RANS) schemes that parameterize the action of all turbulent fluctuations on to the mean flow; and (b) Large Eddy Simulation (LES). The two numerical techniques depict high correlation in terms of droplet deposition at the nasopharynx (as will be discussed through Figure 1). However, it should be noted that while the SST k-*ω* scheme, a 2-equation eddy-viscosity model, is computationally less expensive; it averages the short time-scale flow artifacts, such as the transient vortices (e.g. the low-pressure Dean’s vortices that are common in tortuous channels and can act as droplet attractors); and hence the prediction of droplet transport affected by the simulated ambient airflow may at times contain errors. LES is computationally more expensive, it separates the turbulent flow into large-scale and small-scale motions, and accounts for the small fluctuations through a sub-grid scale model (in this study, Kinetic Energy Transport Model was used as the sub-grid scale model^36^). We took the averaged estimates for regional droplet deposition (along the *in silico* nasal tissue surfaces) from the two schemes, to minimize probable statistical and algorithmic biases.

The computational schemes implemented in the meshed domains employed a segregated solver on ANSYS Fluent, with SIMPLEC pressure-velocity coupling and second-order upwind spatial discretization. Solution convergence was monitored by minimizing the mass continuity and velocity component residuals, and through stabilizing the mass flow rate and static pressure at the airflow outlets. For the pressure-driven flow solutions: typical convergence run-time for a laminar simulation with 5000 iterations was approximately 5–6 hours for 4-processor based parallel computations executed at 4.0 GHz speed. The corresponding run-time for a RANS simulation was *∼* 12 hours; for an LES computation, it was 4–5 days. Note that for the LES work, the simulated flow interval was 0.5 second for the 30 L/min case, with 0.0002 second as the time-step^37^ and it was 0.25 second for the 55 and 85 L/min flow rates with the time-step at 0.0001 second. In the computations, assumed air density was 1.204 kg/m^3^ and 1.825×10^*−*5^ kg/m.s was used as dynamic viscosity of air.

Following set of boundary conditions were enforced during the simulations: (i) zero velocity at the airway-tissue interface i.e. at the walls enclosing the digitized nasal airspace (otherwise commonly referred to as the *no slip* condition), along with “trap” boundary condition for droplets whereby a droplet would come to rest after depositing on the walls; (ii) zero pressure at nostril planes, which were the pressure-inlet zones in the simulations, with “reflect” boundary condition for droplets to mimic the effect of inhalation on the droplet trajectories if they are about to fall out of the anterior nasal domain; and (iii) a negative pressure at the airflow outlet plane, which was the pressure-outlet zone, with “escape” boundary condition for droplets, i.e. allowing for the outgoing droplet trajectories to leave the upper respiratory airspace. Mean inlet-to-outlet pressure gradients were -9.01 Pa at 15 L/min, -26.65 Pa at 30 L/min, -73.73 Pa at 55 L/min, and -155.93 Pa at 85 L/min. For a reference on the general layout of the anatomic regions, see Panel A in Figure 1.

On convergence of the airflow simulations, inhaled droplet dynamics were tracked by Lagrangian-based discrete phase inert particle transport simulations in the ambient airflow; with the localized deposition along the airway walls obtained through numerically integrating transport equations^38^ that consider contribution of the airflow field on the evolution of droplet trajectories, along with the effects for gravity and other body forces such as the Saffman lift force that is exerted by a flow-shear field on small particulates moving transverse to the streamwise direction. Also, the droplet size range is considered large enough to discount Brownian motion effects on their spatial dynamics. Note that the study simulated the transport for 3015 droplets of each size in AR1 and 3000 droplets of each size in AR2, the numbers being same as the number of elements on the nostril inlet planes which were seeded with the to-be-tracked droplets for the droplet transport simulations. For the numerical tracking, the initial mass flow rate of the inert droplets moving normal to the inlet planes into the nasal airspace was required to be non-zero, and was set at 10^*−*20^ kg/s. After the transport simulations, the post-processing of the droplet transmission data along the airway walls provided the regional deposition trends at the nasopharynx.

The numerical methods, discussed and used here, are a significant extension from one of our recent publications^38^ in this journal. The questions explored in the present study are, of course, very different and new, and the findings can be potentially substantial in our evolving field of knowledge on COVID-19. The reader should also note that the numeric protocol has been rigorously validated in the earlier publication^38^, through comparing the regional deposition trends along the inner walls of similar *in silico* nasal anatomic domains to the *in vitro* spray tests performed in 3D-printed solid replicas of the same reconstructions. One may additionally refer to another recent publication^22^ for more details on the digital reconstruction and meshing techniques.

### Estimating virion contamination in respiratory ejecta

Suppose the viral load in a COVID-19 carrier has been assessed to *V* be copies of RNA in each ml of sputum fluid. Let a representative expelled droplet diameter from the carrier be 𝔻 *µ*. With SARS-CoV-2 being a single-stranded RNA virus, the average number of virions embedded in each droplet can then be computed as (*π/*6) *V* 𝔻^3^× 10^*−*12^. Therefore, every 100 droplets of the same size would have (*π/*6) *V* 𝔻^3^ × 10^*−*10^ virions; which, in other words, represents the probability (in %) for a droplet of diameter D *µ*, of containing at least 1 virion.

The study also calculates the number of virions that are depositing at the nasopharynx in unit time. From computational tracking of droplet transport, we can figure out the deposition efficiency of droplets of each size; let the averaged nasopharyngeal deposition efficiency be *η* (in %) for droplets of diameter 𝔻 *µ*. That implies: for every 100 inhaled 𝔻-*µ* droplets, *η* of them are landing on the nasopharynx. Now, if *n* number of such droplets are being ejected by the carrier per minute, then for a closely-positioned individual – the number of 𝔻-*µ* droplets depositing per minute at the nasopharynx is *N* = *n× η/*100. Therefore for a viral load of *𝒱* copies of RNA per ml; the number of virions per minute, that are transmitted to the nasopharynx by the 𝔻-*µ* droplets, is (*πN/*6)*𝒱* 𝔻^3^ × 10^*−*12^.

## Results

### Droplet size range that targets the nasopharynx

The overall droplet size range of 2.5 – 19 *µ* (in AR1: 2.5 – 19 *µ*, in AR2: 2.5 – 15 *µ*) registers the peak, in terms of the percentage of droplets of each size that are deposited at the nasopharynx. The range is determined by a cut-off of at least 5% deposition for around 3000 tracked droplets (viz. 3015 in AR1, 3000 in AR2) of each size. Panel B in Figure 1 displays the heat-maps for nasopharyngeal deposition (NPD) for different droplet sizes, during inhalation at the four tested airflow rates. The discrete droplet sizes, that were tracked, have been marked along the horizontal axis of the heat-maps. The patch bounded by the grey lines can, in fact, be a *definitive graphical technique* to delineate the hazardous droplet size range for various airborne transmissions.

Note that these findings assume that the post-dehydration density of the respiratory droplets (expelled by the carrier and now being inhaled by the exposed individual) is at 1.3 g/ml. If there is little or no dehydration and as such the ejected droplet density remains at *∼* 1 g/ml, the inhaled droplet size range for peak NPD upscales to 3 – 20 *µ* (results available in the online data repository^39^); since the slightly lighter droplets can now penetrate further into the intranasal airspace, the transport process being aided by the ambient inspiratory streamlines.

### Statistical analysis and data interpretation

Panel C in Figure 1 plots the NPD values from RANS (along horizontal axis) and LES (along vertical axis) schemes, implemented for the higher inhalation rates (i.e. 30, 55, and 85 L/min). The simulation outputs are linearly correlated with an average Pearson’s correlation coefficient of 0.98 for 30 L/min, 0.93 for 55 L/min, and 0.95 for 85 L/min. Subsequent check of the slope *m* for the linear best-fit trendline, through the scatter plots of RANS and LES-based NPD data, indicates how similar the estimates are quantitatively; the mean measures therein being *m* = 1.113 for 30 L/min, *m* = 1.052 for 55 L/min, and *m* = 1.177 for 85 L/min; with the value 1 signifying exact equivalence. The statistical operations were carried out on Wolfram Mathematica.

While using a previously reported^7^ ejecta size distribution, the study divides up the percentages for each size bin (i.e. 0–5, 5–10, 10–15 *µ* etc.) uniformly and apportions them to the discrete droplet sizes (belonging to the same size bin) that are tracked (see horizontal axis between the NPD heat-maps in Panel B, Figure 1), to estimate how many droplets of each size would be ejected by the carrier during unit time. The referenced article^7^ described the size bin limits as *A−B*; for consistency, this study interpreted that as droplet sizes (in *µ*) that are *≥ A* and *< B*.

Also at this point, to think of a realistic exposure to a COVID-19 carrier, the vulnerable individual can be considered to inhale at different airflow rates over the duration of exposure. In such context, the Panel A in Figure 2 extracts the averaged nasopharyngeal deposition for the different tested inhalation rates in the two test subjects. Such inhalation-averaged transmission presents an approximate dehydrated droplet size range of 2.5 – 15.0 *µ*, for a minimum 2% NPD for each droplet size.

**Figure 2.**
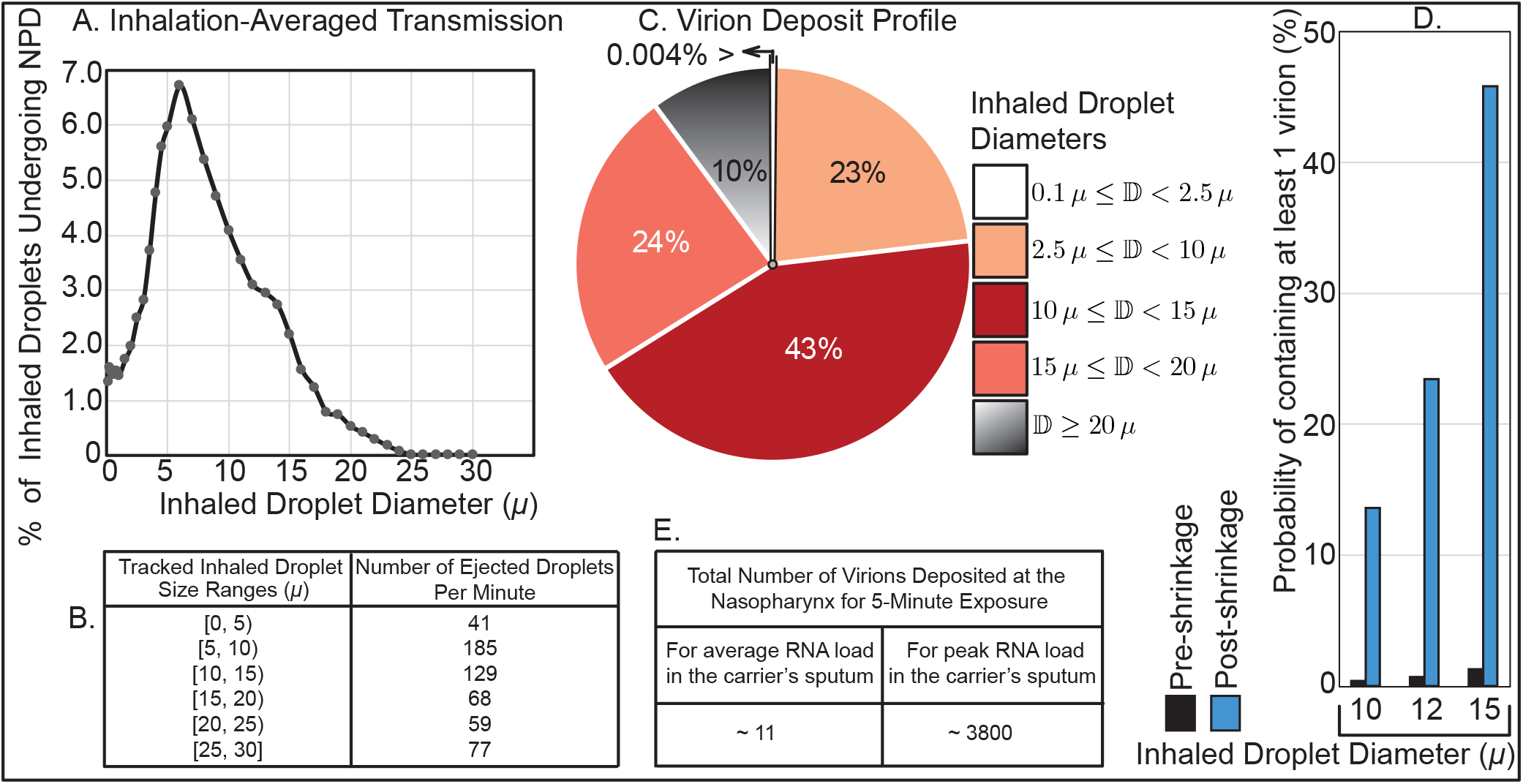
Virion transmission to the nasopharynx: **A**. Graphical representation of the percentage of droplets of each size undergoing deposition at the nasopharynx; averaged from AR1 and AR2, for the tested inhalation rates. **B**. Distribution of droplet sizes ejected each minute during normal speaking, the numbers are calculated from earlier studies on expelled droplet tracking with food coloring^9^. Note the use of parentheses and square brackets to define the size bins; e.g. [*α, β*) implies, as per *set theory* notations, the range of droplet sizes (in *µ*) that are *≥ α* and *< β*. **C**. Pie diagram showing which droplet sizes are dominant contributors for virion transmission at the nasopharynx, for ejecta size distribution as in Panel **B**. Symbol 𝔻 is the inhaled droplet diameter. The numbers assume that the droplets have undergone dehydration before being inhaled into the nasal airspace. **D**. Probabilistic interpretation of a droplet to contain at least 1 virion, based on whether the droplet size at inhalation is pre-dehydration or post-dehydration. **E**. Estimated number of virions that are deposited at the susceptible individual’s nasopharynx via dehydrated inhaled droplets, during close-range exposure to a COVID-19 carrier.

### Droplets that are better at carrying the virions

The next pertinent question is: *how effective are these droplets at carrying virions?* SARS-CoV-2 belongs to a family of single-stranded RNA viruses, and virological assessments^40^ done on the sputum of hospitalized COVID-19 patients show an averaged viral load of 7 ×10^6^ RNA copies/ml of oral fluid, with the peak load being 2.35×10^9^ copies/ml. For the average load, simple calculations (see methods) show that the probability that a dehydrated 10-*µ* droplet (contracted from its original size of *∼* 33 *µ*) will carry at least 1 virion is 13.6%. The same number is 45.8% for a post-shrinkage 15-*µ* droplet. The probability drops exponentially to 0.2% for a 2.5-*µ* dehydrated droplet. Now, with existing data on the size distribution of expelled droplets during normal speaking (see Panel B, Figure 2), the proportion of virion deposits at the nasopharynx by different droplet sizes can be computed (see Panel C, Figure 2) by using the transmission data presented in Figure 1. The deposition trends are again for droplets that are being inhaled post-dehydration.

Conspicuously enough: in the absence of environmental dehydration, the probability of 1 virion being embedded in, for instance, a 10-*µ* droplet plummets to 0.37% (see Panel D, Figure 2). This rationalizes why in geographic regions with high humidity (and hence relatively less dehydration and shrinkage of respiratory ejecta), the pandemic’s spread has been somewhat measured^41,42^.

### What could be COVID-19’s infectious dose

The *infectious dose* is a fundamental virological measure quantifying the number of virions that can go on to start an infection; the value of which is still not conclusively known for SARS-CoV-2^43^. Theoretically, according to the *independent action hypothesis*^44^, even a single virion can potentially establish an infection in highly susceptible systems. Whether the hypothesis is true for humans and specifically for SARS-CoV-2 transmission is as yet undetermined. The rapid spread of COVID-19 though *a priori* suggests a small infective dose for the disease, that is triggering inter-human transmission.

Since it is unethical to expose subjects to SARS-CoV-2 (especially in the absence of well-evidenced remediating therapeutics – as of September 2020), this study introduces a novel strategy synergizing computational tracking and virological data, to quantify the infectious dose. Based on the nasopharyngeal transmission trends (Figure 1) and the virion transmission data (Panels B-C of Figure 2), for a 5-minute exposure: the number of virions depositing at the susceptible individual’s nasopharynx is 11, considering average RNA load in the carrier’s sputum. On the contrary, if the infecting individual is in the disease phase with peak RNA load, as many as 3835 virions will be deposited on the nasopharynx of the exposed individual over 5 minutes (see Panel E in Figure 2).

### A prima facie estimate of infectious dose based on anecdotal reports

To derive a simple *order-of-magnitude* estimate of the SARS-CoV-2 infectious dose, consider the March 2020 Skagit Valley Chorale superspreading incident^5^, where a COVID-19 carrier infected 52 other individuals in a 61-member choir group. Exposure time there was reported to be 2.5 hours. The subjects were situated close to each other; which justifies ignoring the effect of spatial ventilation for a conservative estimate of the number of virions a susceptible individual would have been exposed to. Consequently, for an average RNA load (assuming that the carrier had mild-to-moderate symptoms), the number of virions depositing at a closely-positioned individual’s nasopharynx over that duration approximates to (11*/*5) ×2.5 ×60*≈* 330. So, *∼* 300 can be reckoned as a conservative upper estimate for COVID-19’s infective dose, the order agreeing with preliminary estimates from replication rates of the virus^45^.

One could raise several caveats though; the calculation parameters (especially related to the indoor airflow and ventilation rates, probable spatial fluctuations, and the subject-specific variables associated with the Skagit Valley incident), being used to reach the above estimate for infectious dose, are not, as of now, substantiated by an epidemiological model. The study’s cross-disciplinary strategy (combining numerical simulations of transport in complex anatomic pathways with virological assessments and respiratory ejecta data) could however be potentially used as a sub-component of an epi-model; for an exact quantification of parameters such as the viral infectious dose.

## Discussion

Through tissue culture examinations for respiratory infections, it is fairly well recognized^46^ that only a small fraction of virions are actually able to infect a human cell, and that this fraction decreases rapidly with increasing duration from the time of initial infection of the carrier. So, the SARS-CoV-2 infectivity is being conjectured to peak well before the viral load reaches a maximum. This substantiates the use of averaged viral load in the carrier’s sputum for the virological calculations, while deducing the conservative upper estimate for the SARS-CoV-2 infectious dose.

In this study, whereas the computed data is post-processed to specifically extract the droplet sizes that tend to target the nasopharynx, a vastly larger remainder (comprising predominantly the droplets that are smaller than 5 *µ*) actually go further down the respiratory tract (considering that the air passageways narrow down to just a few microns in the lower airway). However, the significantly larger surface area of the lower airspaces, coupled with the scarcity of ACE2 receptors there, validates the robustness of the modeling framework i.e. focusing on the droplets that deposit on the ACE2-rich epithelial cells at the nasopharynx. Also, the probability of droplets smaller than 5 *µ* to carry a virion is often insignificant; e.g. the probability of containing a virion is only around 1.7% for a 5-*µ* droplet.

Note here that the mathematical approach on the estimation of virion contamination in the respiratory ejecta has, by the very nature of it, presumed a simplistic estimate of viral load in the ejected droplets, based on a continuum-based argument that the spatial distribution of virions could be considered uniform in the sputum. In reality, how the complex rheology of oral fluids might affect the ejecta generation and subsequent break-down, and the resultant volumetric concentration of virions embedded in the expiratory remnants – are also critical open questions.

Finally, this study is limited by the small sample size, primarily owing to the lack of CT scans in subjects with otherwise disease-free airways. To get a realistic insight on the intranasal transport phenomena at the onset of any respiratory infection, it is preferred that we base the *in silico* cavity reconstructions on CT-normal images. Nonetheless, the preliminary findings presented here could be considered an important step in the mechanistic characterization of the transmission dynamics for inhaled pathogens, such as SARS-CoV-2.

### The main takeaways

That the number of virions needed to establish SARS-CoV-2 infection can be of *O* (10^2^) is indeed remarkable! The low order underlines how highly communicable this disease is, especially if discerned in the perspective of the infectious doses for other airborne transmissions, e.g. the infective dose for influenza A virus, when administered through aerosols to human subjects lacking serum neutralizing antibodies, is at least an order greater and ranges between 1950 – 3000 virions^47^.

To summarize: (a) the estimate for the *still-elusive* infectious dose for COVID-19, together with (b) this study’s rigorous detection of the hazardous inhaled droplet sizes (2.5 – 19 *µ*) that specifically target the infection-prone nasopharynx, can provide a key resource in mitigating the pandemic. For example, the information on the droplet sizes that tend to launch the initial infection at the nasopharynx could be utilized to inform public policy on social distancing and in the design of novel masks and face coverings that can screen such droplet sizes and yet be more breathable than the mask respirators that are available now. The findings can also quite significantly provide inputs for the mechanistic design of topical anti-viral therapeutics^48–50^ and targeted intranasal vaccines^51,52^, that would be tailored to land on the infected nasopharynx thereby generating a broader therapeutic window than systemically administered drugs.

## Data Availability

This project has generated simulated quantitative, de-identified data on regional deposition along nasal tissues. The data-sets (including Fluent .cas and .dat files) and the numeric protocols; along with MATLAB codes, Wolfram Mathematica notebooks, and Microsoft Excel spreadsheets used for data post-processing - are available from the corresponding author on request, through a shared-domain Google Drive folder (see the reference list).

https://drive.google.com/drive/folders/1OnEo_fPtSzIqG5_VLKpsg3L5ixG108cI?usp=sharing

## Data availability

This project has generated simulated, quantitative, de-identified data on regional deposition over nasal tissues. The data-sets (including Fluent .cas and .dat files) and the numeric protocols; along with MATLAB codes, Wolfram Mathematica notebooks, and Microsoft Excel spreadsheets used for data post-processing – are available from the corresponding author, through a shared-domain Google Drive folder^39^.

## Funding

This material is based upon work supported by the National Science Foundation (NSF) RAPID Grant 2028069 for COVID-19 research, with the author as the Principal Investigator. Any opinions, findings, and conclusions or recommendations expressed here are, however, those of the author and do not necessarily reflect NSF’s views. Supplemental assistance for the project came from S.B.’s faculty start-up package at South Dakota State University.

## Acknowledgments

Computing facilities at both South Dakota State University and UNC Chapel Hill were used for the work. The author also acknowledges faculty colleagues and mentors Brent A. Senior, MD, FACS, FARS (Professor of Otolaryngology / Neurosurgery at UNC Chapel Hill) and Julia S. Kimbell, PhD (Research Associate Professor at the Department of Otolaryngology / Head and Neck Surgery, UNC Chapel Hill) for fruitful discussions on the findings presented here.

## Conflicts of Interest

The author declares no competing financial interests.

## Additional information

A version of this manuscript has been screened for content and posted on medRxiv. doi: https://doi.org/10.1101/2020.07.27.20162362

## Notes

### Competing Interest Statement

The authors have declared no competing interest.

### Funding Statement

This material is based upon work supported by the National Science Foundation (NSF), under Award Number 2028069 (PI: author S Basu) for COVID-19 research. Any opinions, findings, and conclusions or recommendations expressed here are, however, those of the author and do not necessarily reflect NSF's views. Supplemental assistance for this project came from SB's faculty start-up package at SDSU.

### Author Declarations

Institutional Review Board at the University of North Carolina at Chapel Hill

### Summary of Updates

New details have been added to the Methods section, to clarify the anatomic reconstruction techniques. The sections on results and on discussions have been reviewed and in parts re-drafted to ensure better comprehension by the reader. The reference list has been revised and updated as well.

